# Association between Fat-Infiltrated Axillary Lymph Nodes on Screening Mammography and Cardiometabolic Disease

**DOI:** 10.1101/2021.11.16.21266413

**Authors:** Qingyuan Song, Roberta M. diFlorio-Alexander, Sohum D. Patel, Ryan T. Sieberg, Michael J. Margron, Saif M. Ansari, Margaret R. Karagas, Todd A. Mackenzie, Saeed Hassanpour

## Abstract

**Objective:** Ectopic fat deposition within and around organs is a stronger predictor of cardiometabolic disease status than body mass index. Fat deposition within the lymphatic system is poorly understood. This study examined the association between the prevalence of cardiometabolic disease and ectopic fat deposition within axillary lymph nodes (LNs) visualized on screening mammograms.

**Methods:** A cross-sectional study was conducted on 834 women presenting for full-field digital screening mammography. The status of fat-infiltrated LNs was assessed based on the size and morphology of axillary LNs from screening mammograms. The prevalence of cardiometabolic disease was retrieved from the electronic medical records, including type 2 diabetes mellitus (T2DM), hypertension, dyslipidemia, high blood glucose, cardiovascular disease, stroke, and non-alcoholic fatty liver disease.

**Results:** Fat-infiltrated axillary LNs were associated with a high prevalence of T2DM among all women (adjusted odds ratio: 3.92, 95% CI: [2.40, 6.60], p-value < 0.001) and in subgroups of women with and without obesity. Utilizing the status of fatty LNs improved the classification of T2DM status in addition to age and BMI (1.4% improvement in the area under the receiver operating characteristic curve).

**Conclusion:** Fat-infiltrated axillary LNs visualized on screening mammograms were associated with the prevalence of T2DM. If further validated, fat-infiltrated axillary LNs may represent a novel imaging biomarker of T2DM in women undergoing screening mammography.

## Introduction

Cardiometabolic disease is the leading cause of death among women in the U.S. and is associated with obesity, a condition that now affects greater than 40% of U.S. adults (1–4). Obesity is a heterogeneous condition, and patients with the same body mass index (BMI) have markedly different manifestations of cardiometabolic disease (5). The distribution of fat throughout the body is more closely correlated with cardiometabolic disease than total body fat or BMI(6,7). Existing evidence shows that the obesity phenotype is better characterized by ectopic fat deposition within and around organs that are not designed for excess lipid storage and normally contain small physiologic amounts of adipose tissue (5,8,9). Ectopic adipose within the liver, heart, muscle, and pancreas is associated with organ dysfunction and systemic adverse effects due in part to disturbances in lipid metabolism and chronic low-grade inflammation (5,9–11). A recent position statement from the International Atherosclerosis Society has acknowledged the need to develop tools to assess ectopic fat to better reflect cardiometabolic risk (8). While ectopic fat is a known contributor to cardiometabolic disorders beyond BMI and has been well documented in multiple organ systems (5,9–11), few research studies have examined the association between cardiometabolic disease and ectopic adipose within the immune-lymphatic system.

Lymph nodes (LNs) are small organelles of the immune-lymphatic system distributed throughout the body. On imaging exams, ectopic adipose deposition within benign LNs can be detected as the fat expansion of the radiolucent LN hilum compared to normal LNs that contain only a small quantity of hilar fat (**Figure 1**). In contrast to cortical enlargement seen with reactive nodal hyperplasia and malignant adenopathy, fat-infiltrated nodes are enlarged secondary to adipose deposition within the central hilum with the cortex demonstrating a normal or thinned appearance (12). Recent studies showed that fat-infiltrated axillary LNs on screening mammograms were associated with obesity, independent of age and breast density (10, 12). Fat-infiltrated nodes were also associated with a higher likelihood of axillary metastases in obese women with breast cancer independent of patient and tumor characteristics. Limited studies have demonstrated an association between fat-infiltrated axillary LNs and cardio-metabolic disease. A small study found an association between enlarged fatty nodes and non-alcoholic fatty liver disease (NAFLD) among patients with obesity (13), and a recent study showed that a decrease in the size of fat-infiltrated axillary lymph nodes after bariatric surgery was associated with resolution of dyslipidemia in obese women, independent of weight loss.

**Figure 1.**
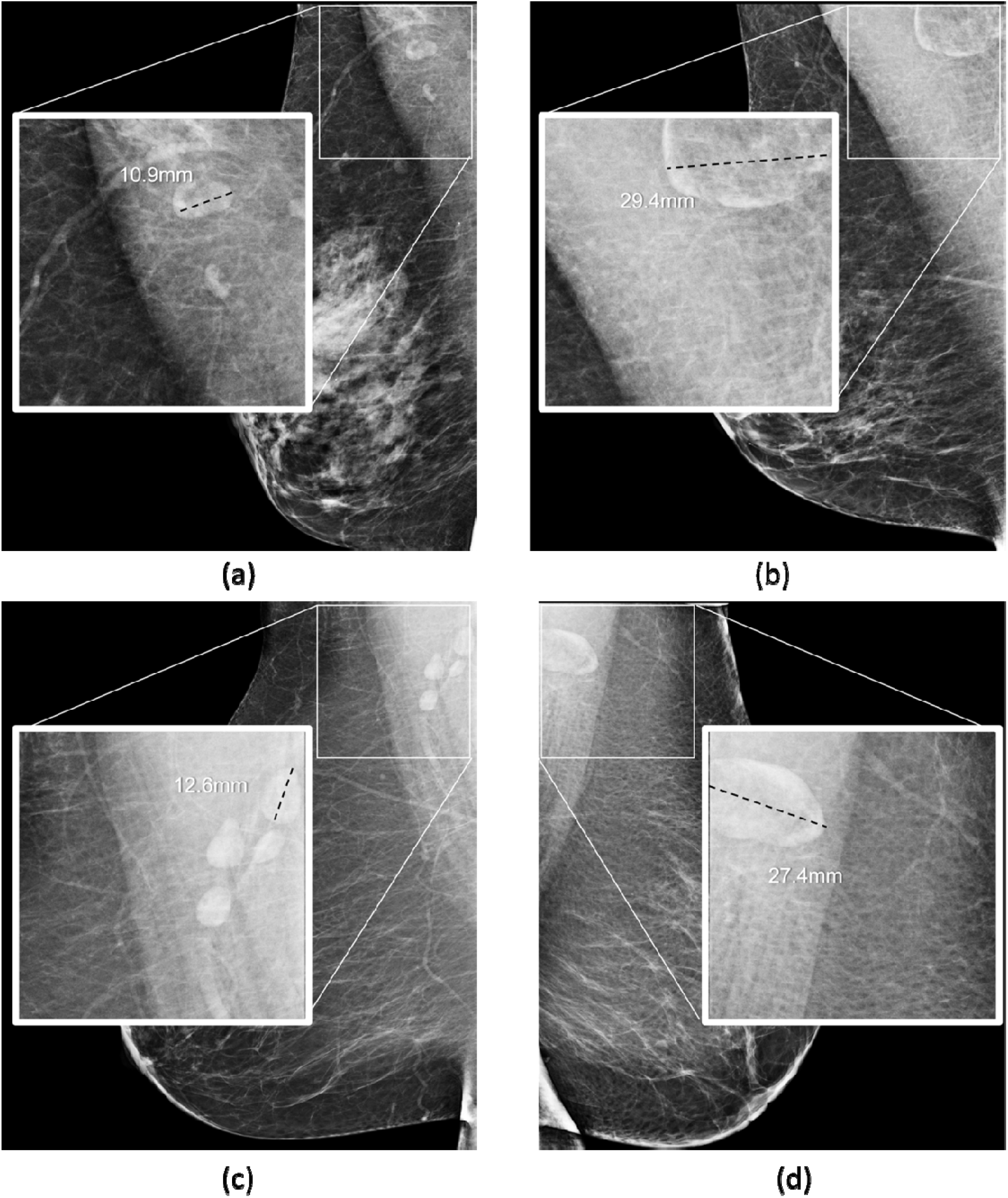
Benign axillary LNs demonstrate various appearances on screening mammography among women with similar age and BMI. a) Female with normal weight (60’s, BMI 22) and a 10.9 mm normal LN; b) Female with normal weight (60’s, BMI 24) and a 29.4 mm fatty LN; c) female with obesity (60’s, BMI 40) and a 12.6 mm normal LN; d) female with obesity (60’s, BMI 40) and a 27.4 mm fatty LN.

Imaging techniques are critical for detecting and quantifying ectopic adipose depots in the body. Many studies have used computerized tomography (CT) and magnetic resonance imaging (MRI) to assess body composition. These studies have demonstrated a strong correlation between ectopic fat and cardiometabolic disease within the liver, muscle, visceral cavity, pancreas, heart, and kidneys, independent of BMI (5,8,9,11). A recent prospective study further demonstrated an increased risk of cardiovascular disease and cancer along with increased ectopic fat among patients with obesity after adjusting for BMI (9). LNs are visible in many imaging studies, including MRI, CT, and ultrasound.

Notably, LNs located in the inferior axilla are also visible on screening mammograms obtained in women without signs or symptoms of breast cancer. Up to 80% of women who undergo a screening mammogram have visible axillary nodes (12,14). Screening mammography, therefore, provides an opportunity to evaluate the appearance of axillary nodes and to further evaluate the association between visible axillary nodes and cardiometabolic disease.

While fat-infiltrated LNs are considered a benign variant of nodal morphology compared to reactive hyperplasia or metastatic adenopathy, the significance of fat-infiltrated LNs in relation to cardiometabolic disease is unknown. We hypothesize that LNs represent a novel ectopic fatty depot and that, similar to ectopic fat seen in other organs, fat-infiltrated nodes are associated with a higher risk of cardiometabolic disease. This study aimed to evaluate the relationship between mammographically visualized fat-infiltrated axillary LNs and features of cardiometabolic diseases, including type 2 diabetes mellitus (T2DM), hypertension, abnormal lipids, high blood glucose, cardiovascular disease (CVD), and NAFLD in women presenting for screening mammography.

## Methods

### Study Group and Image Analysis

This retrospective cohort study included 886 women aged 35 years or older who had a full-field digital mammogram for breast cancer screening at a tertiary medical institution between April 2018 and April 2019. Women presented for screening mammograms without signs or symptoms of breast cancer that would have required diagnostic mammographic evaluation. Participants with negative mammograms classified as Breast Imaging Reporting and Data System-1 (BIRADS-1) were evaluated for axillary LN visualization in the mediolateral oblique (MLO) view by a breast imaging radiologist with 18 years of experience (RDA), as previously described (12). The largest visible axillary LN (index node) was chosen for image analysis by two readers, the breast imaging radiologist (RDA) and a senior radiology resident (RS). Independent measurements of the largest index LN were then obtained along the LNs’ greatest longitudinal axis by both readers, as previously described (12,15). Mammograms were classified as fat-infiltrated if the index LN measured ≥ 18mm, or as normal if the index LN measured < 18mm, using the threshold identified by prior studies (12,15). Cases with discrepancies between the two readers were reviewed and decided by consensus, and the consensus decision was used for the final analysis.

### Collection of Clinical and Cardiometabolic Characteristics

Participants’ clinical characteristics including age and BMI at the time of the exam, and presence of cardiometabolic disease at the date closest to the screening mammogram were obtained from the electronic medical record. If the BMI of a participant could not be found in the record, it was calculated using the formula weight(kg)/[height(m)]^2^. The following cardiometabolic conditions were identified: T2DM, hypertension, abnormal lipids, high blood glucose, CVD, stroke, and NAFLD. The status of each disease was recorded as a binary variable dependent on disease identification in the participants’ problem list. T2DM diagnosis was defined as having a hemoglobin A1C ≥ 6.5% or fasting glucose ≥ 126 mg/dL per the American Diabetes Association definition (16). A diagnosis of hypertension was defined as having a blood pressure greater than 150/90 mmHg for individuals over 60 years old or a blood pressure greater than 140/90 mmHg for those younger than 60 years old according to the Eighth Joint National Committee criteria (17). Abnormal lipids were defined if cholesterol >200 milligrams per deciliter (mg/dL), high-density lipoprotein (HDL) cholesterol < 40 mg/dL, triglycerides > 150 mg/dL, low-density lipoprotein (LDL) >100 mg/dL, or a combination of abnormal serum lipids. Cases with high glucose were identified with blood glucose > 100 mg/dL. A diagnosis of stroke was made in patients with neurologic symptoms corresponding to neuroimaging findings of ischemia. A diagnosis of CVD was defined as any combination of abnormal testing including angiography, stress test (thallium or ECHO), cardiac MRI, or electrocardiogram indicating cardiovascular disease. A diagnosis of NAFLD was made in patients who had a biopsy proven histologic diagnosis of NAFLD or who were diagnosed by liver CT or MRI imaging criteria for NAFLD. Thirty-six of the 886 cases were from outside institutions with no access to the participants’ electronic medical records and were eliminated from the study. Sixteen patients were removed due to a lack of BMI information (or height and weight for BMI calculation). A total of 834 participants were included in this study and the flowchart of the data collection pipeline is shown in **Figure 2**.

**Figure 2.**
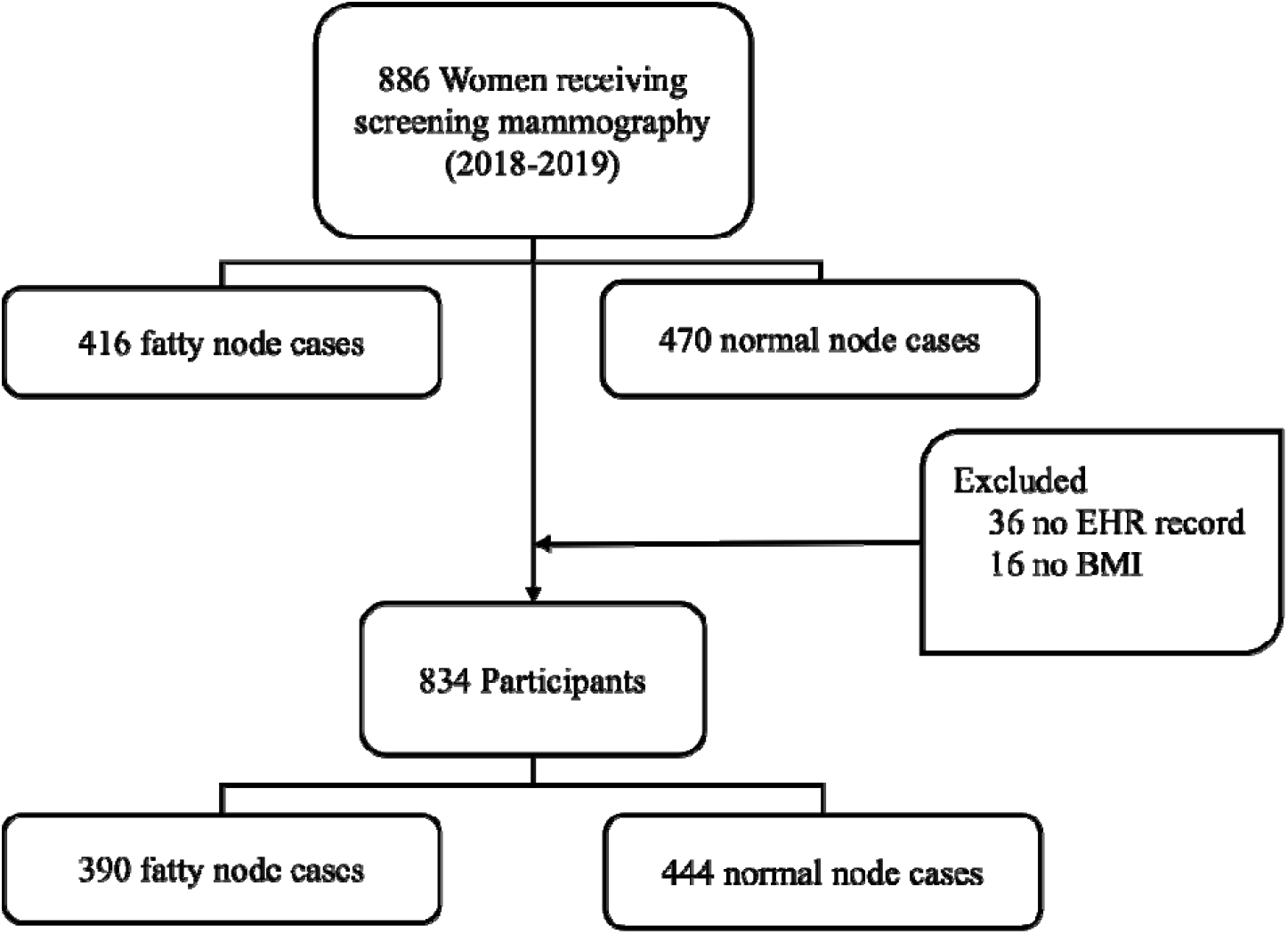
Data collection pipeline of the study participants.

### Statistical Analysis

The association between fatty nodes and cardiometabolic disease was investigated in all individuals and stratified by obesity BMI ≥ 30 kg/m^2^. The crude odds ratios (ORs) between the status of fatty LNs and each cardiometabolic disease were estimated using Fisher’s exact test. The age and BMI-adjusted odds ratios were estimated by multivariate logistic regression models. The 95% confidence interval (CIs) of the odds ratio was calculated for each test and p-value < 0.05 was used as the statistical significance level. An association between cardiometabolic disease and the fat-infiltrated node status was concluded if the estimated odds ratio was greater than 1 with a p-value < 0.05. The statistical analysis was conducted using R version 4.0.3 (R Foundation for Statistical Computing, Vienna, Austria).

### Predicting Cardiometabolic Outcome with Fatty LN and Model Evaluation

Fat-infiltrated LN was evaluated as a potential predictor of cardiometabolic diseases. A baseline model using age and BMI as predictors and a second model incorporating fatty LN status in the predictors were developed to predict each cardiometabolic disease associated with fatty LN. The models’ performances were compared based on their ability to distinguish participants’ cardiometabolic outcome status indicated by the area under the receiver operating characteristic curve (AU-ROC). Bootstrapping on 5-fold cross-validation was performed to avoid overfitting bias. The dataset was randomly split into five folds, each containing 664 training samples (80%) and 166 testing samples (20%). For each fold, 664 samples were randomly sampled from training samples with replacement as the training set. The baseline model and the model using fatty LN as a predictor were trained on the resampled training set, and the testing samples were used to evaluate the models. This process was repeated 2000 times on each fold (10000 times in total) to estimate the distribution of AU-ROC and its difference between predictions with and without fatty LN as a predictor. Similarly, models were evaluated in subgroups of women with and without obesity. The model evaluation workflow was conducted with Python (version 3.6.9, source) and Scikit-learn library (version 0.24, source).

### Ethics

This study is approved by the Institutional Review Board with a waiver of informed consent and is compliant with Health Insurance Portability and Accountability Act (HIPPA).

## Results

The final dataset included 834 female patients with an available problem list in the electronic medical record. Patient age ranged from 35 to 94 years at the date of exam. The distribution of cardiometabolic disease among all women and stratified by obesity is shown in **Figure 3**. The number of patients identified with cardiometabolic disease included 99 T2DM, 306 hypertension, 266 abnormal lipids, 74 high glucose, 72 cardiovascular disease, 11 stroke, and 45 NAFLD (**Figure 3a**). Four patients had type 1 diabetes and were excluded from the analysis of diabetes due to the small population. In this study, 373 (44.7%) patients had BMI 30 kg/m^2^ and were classified as having obesity. Women with obesity were more likely to have cardiometabolic disease and fat-infiltrated nodes. However, 33.5% of women without obesity also had fat-infiltrated nodes identified on screening mammography, and up to 27 % of women without obesity had underlying cardiometabolic disease, most commonly hypertension or dyslipidemia (**Figure 3b**). **Table 1** shows the distribution of patients’ age, BMI, and status of cardiometabolic disease at the time of the screening mammogram among patients with and without fatty nodes with the difference between the two groups indicated by p-value. Fat-infiltrated LNs were found to be more common in older women with a higher BMI. Patients with visible fatty nodes had a higher prevalence of T2DM, hypertension, abnormal lipids, and NAFLD compared to those with normal axillary LNs.

**Figure 3.**
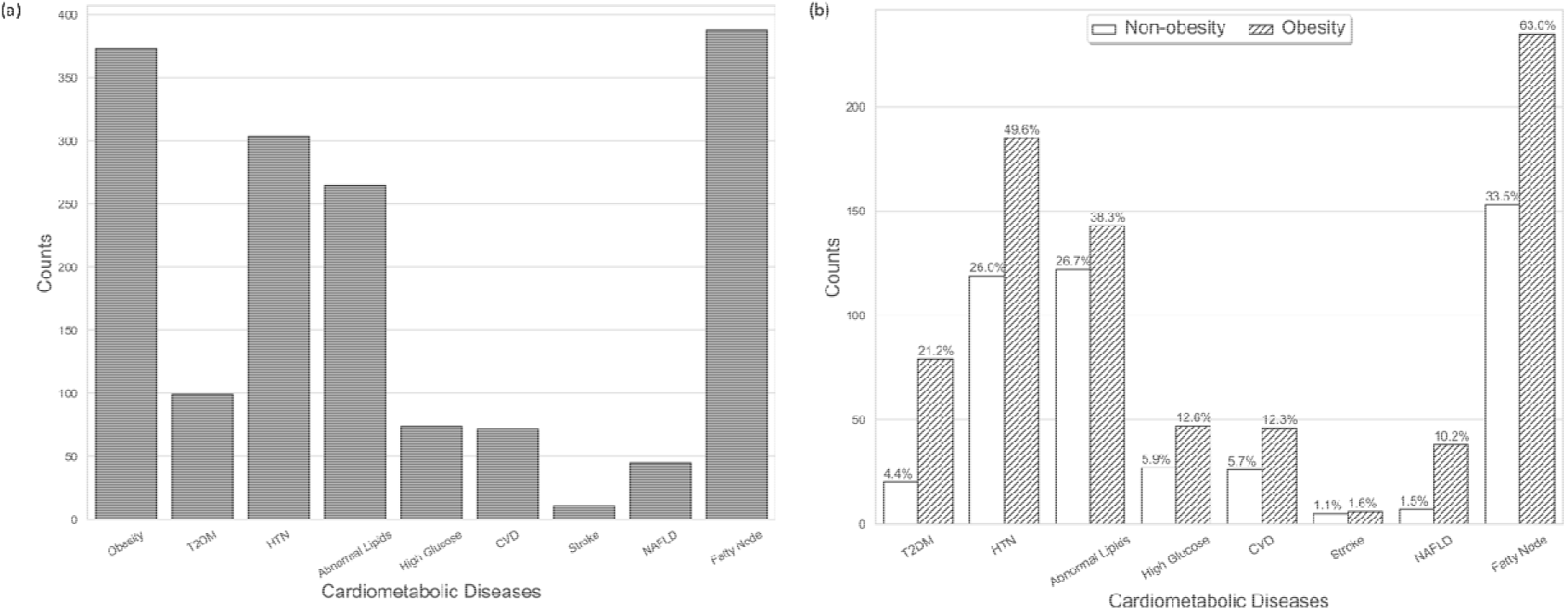
Distribution of cardiometabolic diseases, NAFLD and fatty lymph nodes overall (a) and stratified by obesity status (b).

**Table 1.** Demographics and status of cardiometabolic features of collected cohort, stratified by status of fatty LN.

|  | No fatty node | Fatty node | p-value |
| --- | --- | --- | --- |
| **Total Patients (N, %)** | 444 | 390 |  |
| **Age (mean, SD)** | 61.59 (10.73) | 64.79 (10.00) | < 0.001 |
| **BMI (kg/m<sup>2</sup>) (mean, SD)** | 27.91 (6.56) | 32.57 (7.75) | < 0.001 |
| **Obesity (N, %)** | 138 (31.1) | 235 (60.3) | < 0.001 |
| **Diabetes (N, %)** |  |  | < 0.001 |
| **Type 1** | 2 (0.5) | 2 (0.5) |  |
| **Type 2** | 25 (5.6) | 74 (19.0) | < 0.001 |
| **Hypertension (N, %)** | 130 (29.3) | 176 (45.1) | < 0.001 |
| **Abnormal lipids (N, %)** | 119 (26.8) | 147 (37.7) | 0.001 |
| **High glucose (N, %)** | 37 (8.3) | 37 (9.5) | 0.644 |
| **CVD (N, %)** | 32 (7.2) | 40 (10.3) | 0.150 |
| **Stroke (N, %)** | 6 (1.4) | 5 (1.3) | 1.000 |
| **NAFLD (N, %)** | 15 (3.4) | 30 (7.7) | 0.009 |

### Association between Fat-enlarged LNs and Cardiometabolic Disease

**Figure 4** describes the crude OR between fatty LNs and each cardiometabolic disease based on Fisher’s exact test, and the age and BMI-adjusted ORs estimated from logistic regression, along with the corresponding p-values. The crude OR estimates showed fatty node was associated with several cardiometabolic disease, including T2DM (crude OR = 3.92, p < 0.001), hypertension (crude OR = 1.98, p < 0.001), abnormal lipids (crude OR = 1.65, p < 0.001), and NAFLD (crude OR = 2.38, p = 0.008). However, after adjusting for age and BMI, only T2DM remained significantly associated with fat-infiltrated nodes (adjusted OR = 2.45, p < 0.001), while age and/or BMI were significant confounders for hypertension, abnormal lipids, and NAFLD. The detailed results for unadjusted and age and BMI-adjusted analyses are shown in **Supplementary Table 1**.

**Figure 4.**
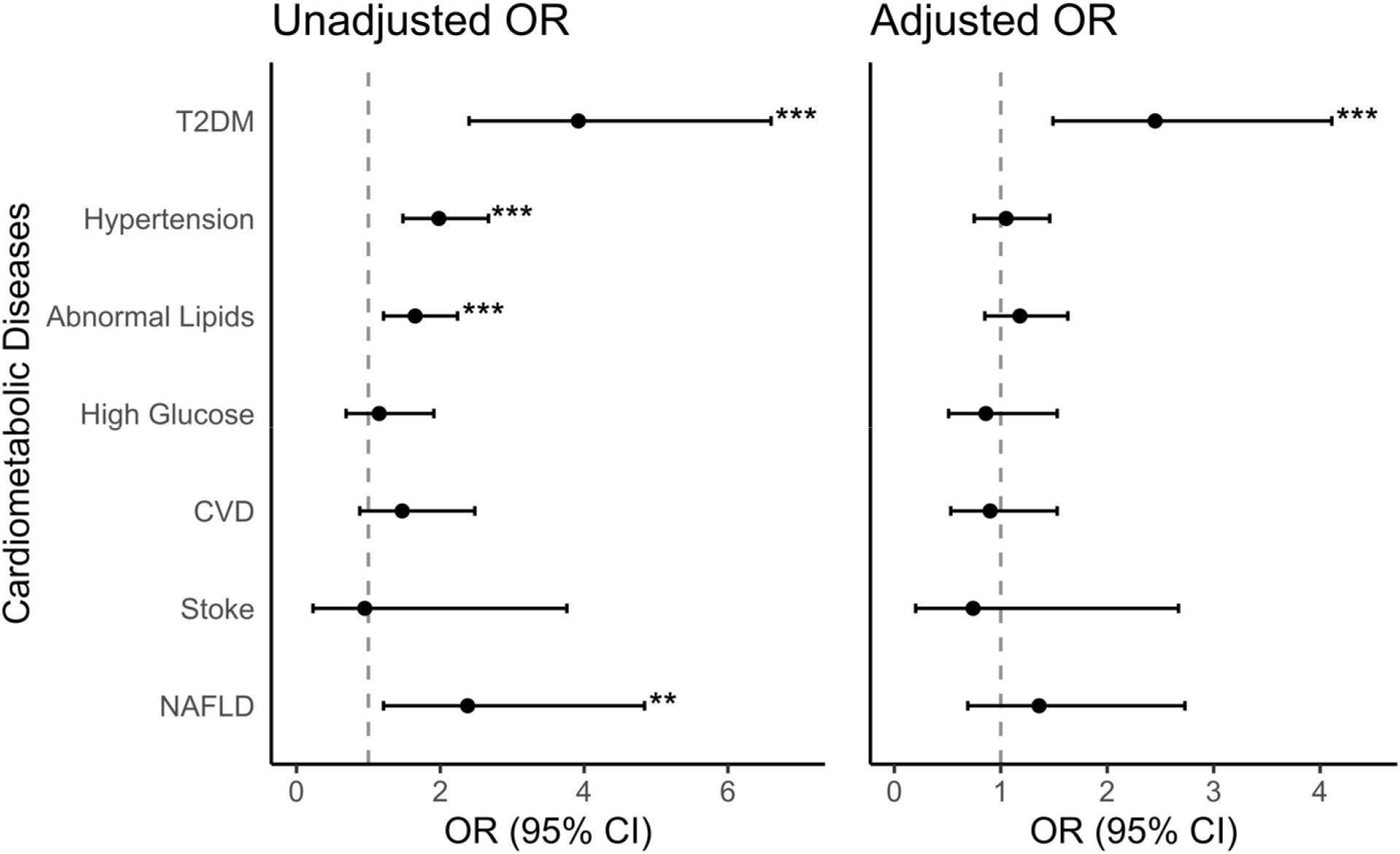
Association between fat-enlarged axillary LNs and collected cardiometabolic disease status. Data are presented as unadjusted, and age and BMI-adjusted odds ratio estimates with 95% CI between binary fatty nodes status and binary cardiometabolic disease status of all patients in our cohort. T2DM: type 2 diabetes mellitus; CVD: cardiovascular disease; NAFLD: non-alcoholic fatty liver disease. *** p < 0.001 ** p < 0.01 * p < 0.05

The estimated odds ratio between stroke and fatty LNs was excluded from the analysis of subgroups with and without obesity due to a small number of patients with stroke. T2DM was linked to fatty LNs in both groups with and without obesity, according to findings of the subgroup analysis. The estimated OR was higher in the group without obesity (adjusted OR = 2.99, p = 0.029) compared to the group with obesity (adjusted OR = 2.03, p = 0.018) although the confidence intervals overlapped. The detailed results for obesity-stratified analysis are shown in **Supplementary Table 2**.

### Performance of T2DM Prediction with Fatty LNs Status

The model evaluation indicated an overall improved T2DM prediction with fat-infiltrated LN status. The LOOCV AU-ROC increased from 0.758 with the baseline model to 0.772 for all women when fatty LN status was included in the predictors. Adding fatty LN status to the model improved the model’s discriminating capacity by 0.014, 0.023 and 0.033 in all participants, and those with and without obesity, respectively. The detailed model performance of each model is shown in **Table 2**.

**Table 2.** Comparison of AU-ROC with standard error (SE) between the baseline model (**Model 1**) and the model including fat-infiltrated LNs (**Model 2**) for T2DM prediction.

| | AU-ROC ± SE | | Difference ± SE |
| --- | --- | --- | --- |
|  | Model 1 | Model 2 |  |
| **All** | 0.758±0.0006 | 0.772±0.0006 | 0.014±0.0003 |
| **With Obesity** | 0.625±0.0008 | 0.648±0.0007 | 0.023±0.0006 |
| **Without Obesity** | 0.646±0.0011 | 0.679±0.0009 | 0.033±0.0009 |

## Discussion

This study examined the association between cardiometabolic disease and fat-infiltrated axillary LNs visualized on screening mammograms of adult women. After adjusting for age and BMI, there was a significant association between T2DM and fat-infiltrated axillary LNs. This significant association was maintained between T2DM and fat-infiltrated LNs in subgroups of women with and without obesity. Fat-infiltrated LNs were also associated with hypertension, abnormal lipids, and NAFLD in the crude analysis; however, this association did not remain significant when adjusted for age and BMI. The evaluation of models classifying T2DM status showed that adding fatty LN status to the model improved the classification of T2DM in women with and without obesity. The above results support the possibility that fat-infiltrated LNs may represent a novel ectopic fat depot associated with cardiometabolic disease, similar to the association between other ectopic fat depots and cardiometabolic disease.

Obesity is a risk factor for cardiometabolic disease; however, the obesity phenotype is heterogeneous and cardiometabolic risk is not accurately represented by BMI. In contrast, ectopic adiposity has been shown to be a better predictor of cardiometabolic disease than BMI. Multiple studies have demonstrated an association between ectopic visceral fat and T2DM independent of BMI (18). The Dallas Heart Study showed that ectopic visceral fat was independentlassociated with T2DM among adults with obesity, while BMI, total body fat, and abdominal subcutaneous fat were not associated with incident diabetes mellitus (19,20). Patients with NAFLD have been shown to have a higher risk of T2DM irrespective of BMI (21). A recent longitudinal study of nearly 16,000 subjects showed that ectopic fat had a markedly elevated risk for developing T2DM compared to BMI-defined obesity (21). In addition, epicardial adipose deposition is associated with insulin resistance and T2DM in adults with and without obesity (23). The current study’s findings are in keeping with the previous studies. It suggested that ectopic LN fat may be an indicator of T2DM among groups of women with and without obesity, independent of BMI.

Fatty nodes are considered a benign anatomic variant in contrast to metastatic or reactive lymphadenopathy. However, a large body of evidence has shown that fat is not inert. Unlike fat distributed in the classic subcutaneous compartment, ectopic fat is associated with chronic inflammation, dysregulated lipid metabolism, and altered adipokine secretion that may lead to local organ dysfunction and systemic adverse effects (5,9–11,19,20,24). Proposed mechanisms accounting for adverse health effects associated with ectopic fat deposition in other organs may similarly explain the association between ectopic fat-infiltrated LNs and diabetes found in the current study. A small physiologic quantity of hilar nodal fat may be beneficial in protecting traversing vessels and lymphatics as they course through the nodal hilum. However, a large volume of ectopic hilar fat may have detrimental effects and impair nodal function by compressing hilar structures, such as low resistance venous and lymphatic channels. This mechanism is analogous to the adverse effect linked to kidney dysfunction caused by ectopic adipose deposition in the central renal sinus (25). LNs are critical organs for lipid trafficking, and decreased flow through the efferent lymphatics due to fat compression may alter lipid transportation and negatively impact insulin resistance. Finally, the mass effect of the fat-expanded hilum on the adjacent LN cortex may negatively affect cortical function, as supported by studies showing decreased immune cell function in obese mice (26,27). The adverse health impact of ectopic fat in other organs has also been linked to poor cancer outcomes. A recent study showing a higher likelihood of nodal metastases in obese women with fat enlarged axillary nodes further supports our hypothesis that ectopic nodal fat may negatively impact LN function (15).

This study showed that ectopic fat in axillary LNs identified on screening mammograms is significantly associated with T2DM in women with and without obesity, independent of BMI and age. Imaging studies contain data beyond their intended use. The expanding field of body composition research has characterized imaging features of adverse health outcomes on imaging exams obtained for other indications (28,29). Body composition studies have been at the forefront of research evaluating ectopic fat deposition, and many studies demonstrated a strong correlation between poor cardiometabolic status and ectopic adipose within the visceral cavity, liver, and muscle (5,8,9,11). It has been proposed that in this era of precision medicine, cardiovascular risk assessment among people with obesity should be refined by measures of ectopic fat deposition rather than BMI (5,8,29,30). The majority of women in the United States undergo mammography screening but do not undergo screening for cardiovascular disease (14). This makes opportunistic axillary LN evaluation on screening mammograms an ideal target for future research assessing the association between fat-infiltrated LNs and cardiometabolic disease.

The current study was the first to examine the association between fat deposition in the immune system and cardiometabolic disease status. The strengths of this pilot study include a significant sample size and the exploration of a novel concept that fat-infiltrated axillary nodes may represent an ectopic fatty depot. Like other ectopic fatty depots, fat-infiltrated axillary nodes were significantly associated with T2DM. This study had several limitations. First, the prevalence of cardiometabolic disease may be underestimated in this study as the conditions determined from the problem list may under-report or fail to update patient health status, which might result in a smaller effect size. Additional studies are needed to further investigate the findings of the current study. Second, this pilot study included women from a single institution with visible LNs on their screening mammograms. Variable LN visibility is due to the differences in patient body habitus and technical differences related to patient positioning at the time of the exam. No appreciable differences have been reported among women with and without visible LNs (15,29). Nonetheless, inconsistent mammographic visualization of axillary LNs may introduce bias when evaluating the association between fatty nodes and cardiometabolic disease status. Finally, the current study used a cross-sectional design and did not evaluate the causal relationship between fatty LN and cardiometabolic disease. Prospective studies are needed to investigate the potential mechanisms contributing to the association between ectopic nodal fat and cardiometabolic disease. Despite variable mammographic LN visualization, the ability to leverage an existing screening study to opportunistically measure ectopic LN fat makes this opportunistic technique potentially scalable. The current study confirms findings in prior studies showing a strong association between fatty nodes and obesity and further suggests that fat-infiltrated LNs may represent a novel ectopic fat depot. If confirmed in larger multi-site studies, the ability to provide cardiometabolic risk assessment by leveraging a highly utilized breast cancer screening exam could have a significant clinical impact without added cost. Opportunistic mammographic axillary LN evaluation is therefore an ideal target for future research investigating the association between fat-infiltrated LNs and cardiometabolic disease.

## Conclusion

In conclusion, a significant association between fat-infiltrated axillary LNs and T2DM was found in 834 women with visible axillary LNs on screening mammograms independent of age and BMI. The identification of mammographic fat-infiltrated axillary LNs could potentially improve the assessment of T2DM.

## Data Availability

All data produced in the present study are available upon reasonable request to the authors.

