## Supplementary Table 1 for "Association between Fat-Infiltrated Axillary Lymph Nodes on Screening Mammography and Cardiometabolic Disease"

**Table S1.** Crude and adjusted OR and 95% confidence intervals of relation between fatty nodes and the status of cardiometabolic disease in all collected patients.

|  | **Crude OR (95% CI)** | **p-value** | **Adjusted OR (95% CI)** | **p-value** |
| --- | --- | --- | --- | --- |
| **Type 2 Diabetes** | **3.92 (2.40, 6.60)** | **<0.001** | **2.45 (1.49, 4.11)** | **< 0.001** |
| **Hypertension** | **1.98 (1.48, 2.67)** | **< 0.001** | 1.05 (0.75, 1.46) | 0.766 |
| **Abnormal Lipids** | **1.65 (1.21, 2.24)** | **<0.001** | 1.18 (0.85, 1.63) | 0.312 |
| **High glucose** | 1.15 (0.69, 1.91) | 0.626 | 0.86 (0.51, 1.53) | 0.558 |
| **CVD** | 1.47 (0.88, 2.48) | 0.138 | 0.90 (0.53, 1.53) | 0.705 |
| **Stroke** | 0.95 (0.23, 3.76) | 1.000 | 0.74 (0.20, 2.67) | 0.647 |
| **NAFLD** | **2.38 (1.21, 4.84)** | **0.008** | 1.36 (0.69, 2.73) | 0.382 |
