## Supplementary Table 2 for "Association between Fat-Infiltrated Axillary Lymph Nodes on Screening Mammography and Cardiometabolic Disease"

**Table S2.** Adjusted odds ratio and 95% confidence intervals of relation between fatty nodes and status of cardiometabolic disease, stratified for status of obesity (BMI threshold = 30).

|  | **Obese (BMI >= 30, N = 346)** | | **Non-obese (BMI < 30, N = 436)** | |
| --- | --- | --- | --- | --- |
|  | **Adjusted odds ratio (95% CI)** | **p-value** | **Adjusted odds ratio (95% CI)** | **p-value** |
| **Type 2 Diabetes** | **2.03 (1.15, 3.74)** | **0.018** | **2.99 (1.15, 8.40)** | **0.029** |
| **Hypertension** | 1.01 (0.63, 1.62) | 0.951 | 1.06 (0.66, 1.69) | 0.810 |
| **Dyslipidemia** | 0.95 (0.61, 1.51) | 0.849 | 1.32 (0.83, 2.09) | 0.242 |
| **High glucose** | 0.86 (0.46, 1.65) | 0.652 | 0.66 (0.27, 1.52) | 0.343 |
| **Cardiovascular diseases** | 0.79 (0.41, 1.56) | 0.492 | 1.05 (0.44, 2.44) | 0.903 |
| **Stroke** | 2.69 (0.42, 52.38) | 0.372 | - | - |
| **NAFLD** | 1.49 (0.71, 3.36) | 0.315 | 1.36 (0.69, 2.73) | 0.382 |
